# Comparative Effectiveness of TTR Stabilizers for the Treatment of ATTR-CM Using Real-World Evidence: Acoramidis versus Tafamidis

**DOI:** 10.64898/2026.04.24.26351684

**Authors:** Richard Wright, Trejeeve Martyn, Allison Keshishian, Elizabeth Nagelhout, Bret Zeldow, Margarita Udall, David Lanfear, Daniel P. Judge

**Affiliations:** Pacific Heart Institute, Los Angeles, CA, USA; Department of Cardiovascular Medicine, Heart, Vascular and Thoracic Institute, Cleveland Clinic, Cleveland, OH, USA; Genesis Research Group, Hoboken, NJ, USA; BridgeBio Pharma, Inc., San Francisco, CA, USA; Department of Medicine, Henry Ford Health, Detroit, MI, USA; Department of Medicine, Medical University of South Carolina, Charleston, SC, USA

## Abstract

**Background:** Progression of transthyretin (TTR) amyloid cardiomyopathy (ATTR-CM) can lead to worsening congestion requiring diuretic intensification (DI), heart failure (HF)-related hospitalizations (HFH), and death. Tafamidis was the only approved ATTR-CM therapy in the US from 2019 until the 2024 approval of acoramidis, which achieves near-complete (≥90%) TTR stabilization. As head-to-head trials are lacking, real-world comparative effectiveness (CE) data are needed to guide treatment selection.

**Objective:** To evaluate real-world CE of acoramidis versus tafamidis in newly treated patients with ATTR-CM.

**Methods:** Retrospective study using Komodo Healthcare Map^®^ US claims data tokenized to Claritas. Patients newly initiating acoramidis or tafamidis between 12/11/2024 and 04/30/2025 with ≥1 prescription claim (first defined as index date) and ≥6 months of continuous enrollment preindex date were included and followed until disenrollment, death, treatment switch, or study end date (07/31/2025). Outcomes included DI (initiation or dose-equivalent escalation of oral loop diuretics, parenteral loop diuretic use, or addition of thiazide-like diuretic) and a composite of DI, HFH (inpatient admission with a HF-related ICD-10-CM diagnosis code in any position), and mortality. Propensity score weighting balanced baseline characteristics, disease severity, comorbidity burden, and baseline medication use. Time-to-event outcomes were assessed using weighted Cox proportional hazards models.

**Results:** After weighting, acoramidis (n=170) and tafamidis (weighted sample size=448) patients were comparable at baseline (mean age, 78.6 vs 78.7 years; male, 80.0% vs 80.2%) with mean follow-up of 139 and 143 days, respectively. DI cumulative incidence curves separated early and remained divergent, with acoramidis significantly reducing the hazard of DI events by 43% compared with tafamidis (11.8% vs 20.5%; HR, 0.57; 95% CI, 0.35-0.92; *P*=0.021). Acoramidis also had a significantly lower risk of composite events, with a 34% reduction in hazard compared with tafamidis (17.6% vs 26.4%; HR, 0.66; 95% CI, 0.44-0.99; *P*=0.046).

**Conclusions:** In this first real-world CE study of newly treated patients, acoramidis had significantly lower risk of DI events and composite events of DI, HFH, and mortality than tafamidis, potentially supporting improved clinical stability with acoramidis initiation. Additional evaluation with longer follow-up, larger cohorts, and/or prospective clinical outcomes is warranted.

## Introduction

Transthyretin (TTR) amyloid cardiomyopathy (ATTR-CM) is an infiltrative disease caused by unstable TTR that forms amyloid fibrils, which deposit within the myocardium.^1,2^ Disease progression often begins with congestion requiring diuretic treatment, followed by heart failure (HF)-related hospitalization (HFH), which increases mortality risk.^1,3^ Outpatient diuretic intensification (DI) is associated with increased mortality and is viewed as an early marker of disease progression.^3-5^ Upon its approval in 2019, tafamidis, a TTR stabilizer, was the only therapy for ATTR-CM until the 2024 approval of acoramidis, which achieves near-complete (≥90%) TTR stabilization.^2,6,7^ Both stabilizers reduce mortality and cardiovascular-related hospitalization compared with placebo.^6,7^ In the absence of head-to-head randomized trials, real-world comparative effectiveness data are needed to inform clinical decision-making and optimize patient outcomes. We evaluated the real-world effectiveness of acoramidis versus tafamidis in newly treated patients with ATTR-CM, using DI as an early, clinically meaningful outcome plus a composite of DI, HFH, and mortality.

## Methods

This was a retrospective longitudinal new-user, active-comparator claims cohort analysis of US patients from the Komodo Healthcare Map^®^, with acoramidis data tokenized with Claritas (a data aggregator, which consolidates prescription data between specialty pharmacies and the acoramidis manufacturer’s hub) via Datavant token. The Komodo database complies with the Health Insurance Portability and Accountability Act. Given the deidentified nature of the data, patient consent and institutional review board approval were not required. Aggregate cohort-level results were reported. This study followed the International Society for Pharmacoeconomics and Outcomes Research (ISPOR) guidelines for comparative effectiveness research.^8^

Patients were included if they were aged ≥18 years with ≥1 claim for acoramidis or tafamidis during the identification period from 12/11/2024 to 4/30/2025 and had ≥6 months of continuous enrollment before the index date (first acoramidis or tafamidis prescription). Patients were excluded if they had multiple myeloma, light chain amyloidosis, hematopoietic stem cell treatment at any time during the study period, had evidence of prior treatment with tafamidis, acoramidis, or a TTR silencer before the index date, or >1 ATTR-CM treatment on index date. Patients were followed until disenrollment, death, treatment switch, or study end date (7/31/2025).

The primary outcome was DI, defined as initiation or dose-equivalent escalation of oral loop diuretics, parenteral loop diuretics use, or addition of a thiazide-type diuretic. The secondary outcome was a composite of DI, HFH (inpatient admission with an HF-related ICD-10-CM diagnosis code in any position), and mortality. A subanalysis of oral loop DI was also conducted.

The statistical analysis plan followed the ISPOR guidelines,^8^ including the specification of the research question, prespecified analytical plans such as eligibility criteria, potential biases, outcome measures, handling of missing data, and all methods described herein. Categorical variables were summarized by frequency and percentage, and continuous variables by mean and SD. Propensity score weighting was used to balance baseline characteristics, time from diagnosis, measures of disease severity, comorbidity burden and medication use, including loop diuretics (**Table 1)**. Collinearity was evaluated using variance inflation factors, and for highly correlated predictors, variables with greater clinical relevance and more robust data were retained. Logistic regression estimated the probability of receiving acoramidis based on prespecified covariates, and adequate balance was defined a priori as absolute standardized mean differences <0.1 for all covariates. Time-to-event outcomes were assessed using weighted Cox proportional hazards models for DI and oral loop DI separately and then for the first of any component of the composite, with tafamidis as the reference group. Kaplan-Meier estimates were used to assess the cumulative incidence of each outcome. Statistical significance was defined as two-sided *P*<0.05. We assessed falsification outcomes (any fracture, pneumonia, urinary tract infection, vaccination) to detect residual bias.

**Table 1.** Baseline patient demographics and clinical characteristics before and after propensity score weighting.

| Characteristic | Unweighted |  | Weighted |  |
| --- | --- | --- | --- | --- |
|  | Acoramidis<br>(n = 175) | Tafamidis<br>(n = 697) | Acoramidis<br>(n = 170) | Tafamidis<br>(WSS = 448) |
| **Age, mean (SD), y** | 78.7 (7.7) | 80.0 (6.9) | 78.6 (7.7) | 78.7 (7.3) |
| **Age group, n (%)<sup>a</sup>** |  |  |  |  |
| <80 y | 87 (49.7) | 285 (40.9) | 84 (49.4) | 224 (49.9) |
| ≥80 y | 88 (50.3) | 412 (59.1) | 86 (50.6) | 224 (50.1) |
| **Sex, n (%)<sup>a</sup>** |  |  |  |  |
| Male | 136 (77.7) | 526 (75.5) | 136 (80.0) | 359 (80.2) |
| Female | 34 (19.4) | 149 (21.4) | 34 (20.0) | 89 (19.8) |
| Unknown | 5 (2.9) | 22 (3.2) | NA<sup>b</sup> | NA<sup>b</sup> |
| **Race, n (%)** |  |  |  |  |
| White<sup>a</sup> | 104 (59.4) | 434 (62.3) | 102 (60.0) | 268 (59.9) |
| Black or African American | 48 (27.4) | 182 (26.1) | 45 (26.5) | 121 (27.0) |
| Other<sup>a</sup> | 16 (9.1) | 66 (9.5) | 16 (9.4) | 51 (11.3) |
| Missing/Unknown | 7 (4.0) | 15 (2.2) | 7 (4.1) | 8 (1.8) |
| **Region, n (%)<sup>a</sup>** |  |  |  |  |
| Northeast | 45 (25.7) | 209 (30.0) | 42 (24.7) | 108 (24.1) |
| Midwest | 38 (21.7) | 158 (22.7) | 38 (22.4) | 100 (22.3) |
| South | 63 (36.0) | 217 (31.1) | 61 (35.9) | 164 (36.6) |
| West | 29 (16.6) | 109 (15.6) | 29 (17.1) | 76 (17.0) |
| Missing/Unknown | 0 | 4 (0.6) | NA<sup>b</sup> | NA<sup>b</sup> |
| **Insurance type, n (%)** |  |  |  |  |
| Medicare | 149 (85.1) | 637 (91.4) | 144 (84.7) | 403 (89.9) |
| Commercial | 13 (7.4) | 19 (2.7) | 13 (7.6) | 17 (3.8) |
| Other | 11 (6.3) | 29 (4.2) | 11 (6.5) | 21 (4.7) |
| Missing/Unknown | 2 (1.1) | 12 (1.7) | 2 (1.2) | 7 (1.6) |
| **Year of ATTR-CM diagnosis, n (%)<sup>a</sup>** |  |  |  |  |
| 2019-2023 | 23 (13.1) | 273 (39.2) | 23 (13.5) | 60 (13.4) |
| 2024-2025 | 140 (80.0) | 397 (57.0) | 135 (79.4) | 355 (79.2) |
| Unknown | 12 (6.9) | 27 (3.9) | 12 (7.1) | 33 (7.3) |
| **Medication use before or on index, n (%)** |  |  |  |  |
| Beta blocker | 112 (64.0) | 427 (61.3) | 109 (64.1) | 288 (64.3) |
| Mineralocorticoid receptor antagonist | 62 (35.4) | 270 (38.7) | 61 (35.9) | 175 (39.0) |
| Sodium-glucose cotransporter 2 inhibitor<sup>a</sup> | 75 (42.9) | 271 (38.9) | 73 (42.9) | 197 (44.0) |
| **Loop diuretic dose at acoramidis or tafamidis initiation, n (%)<sup>a,c</sup>** |  |  |  |  |
| No loop diuretic dose | 79 (45.1) | 254 (36.4) | 76 (44.7) | 200 (44.7) |
| <60 mg furosemide | 78 (44.6) | 316 (45.3) | 76 (44.7) | 199 (44.5) |
| ≥60 mg furosemide | 18 (10.3) | 127 (18.2) | 18 (10.6) | 48 (10.8) |
| **Charlson Comorbidity Index score, mean (SD)<sup>a</sup>** | 3.7 (2.2) | 3.6 (2.3) | 3.7 (2.2) | 3.7 (2.3) |
| **Comorbidities, n (%)<sup>a</sup>** |  |  |  |  |
| Atrial fibrillation | 100 (57.1) | 429 (61.5) | 96 (56.5) | 253 (56.5) |
| Chronic kidney disease | 81 (46.3) | 344 (49.4) | 77 (45.3) | 209 (46.6) |
| Hypertension | 147 (84.0) | 574 (82.4) | 142 (83.5) | 375 (83.6) |
| Pericarditis | 21 (12.0) | 49 (7.0) | 19 (11.2) | 51 (11.3) |
| **CV-related hospitalization within 6 months of index, n (%)<sup>a</sup>** | 45 (25.7) | 148 (21.2) | 43 (25.3) | 116 (25.8) |
| **HFH within 6 months of index, n (%)** | 40 (22.9) | 131 (18.8) | 38 (22.4) | 99 (22.0) |
| **ED visit within 6 months of index, n (%)<sup>a</sup>** | 61 (34.9) | 212 (30.4) | 61 (35.9) | 160 (35.7) |

<sup>a</sup>Characteristics used in propensity score weighting.
<sup>b</sup>Patients of unknown sex or region were removed for modeling purposes.
<sup>c</sup>Loop diuretic dose was measured closest to the index date within 90 days before or 7 days after index date.
ATTR-CM, transthyretin amyloid cardiomyopathy; CV, cardiovascular; ED, emergency department; HFH, heart failure-related hospitalization; WSS, weighted sample size.

## Results

Overall, 175 and 697 patients newly treated with acoramidis and tafamidis, respectively, met all inclusion/exclusion criteria. After weighting, acoramidis (n=170) and tafamidis (weighted sample size=448) patients were comparable at baseline (**Table 1**). Mean days of follow-up were 139 and 143 for acoramidis and tafamidis, respectively.

Cumulative incidence curves for the first event of DI **(Figure 1A**), the composite (**Figure 1B**), and oral loop DI (**Figure 1C**) separated within weeks of treatment initiation and remained divergent over time. Acoramidis was associated with significantly reduced DI events (11.8% vs 20.5%; hazard ratio [HR], 0.57; 95% CI, 0.35-0.92; *P*=0.021), composite events (17.6% vs 26.4%; HR, 0.66; 95% CI, 0.44-0.99; *P*=0.046), and oral loop DI events (10.0% vs 18.9%; HR, 0.52; 95% CI, 0.31-0.87; *P*=0.014) compared with tafamidis. The proportion of patients experiencing HFH was 10.6% versus 12.3% (HR, 0.90; 95% CI, 0.53-1.55; *P*=0.71) and experiencing mortality was 1.2% versus 2.3% (HR, 0.52; 95% CI, 0.11-2.42; *P*=0.41) for acoramidis and tafamidis, respectively. All falsification outcomes were nonsignificant.

**Figure 1.**
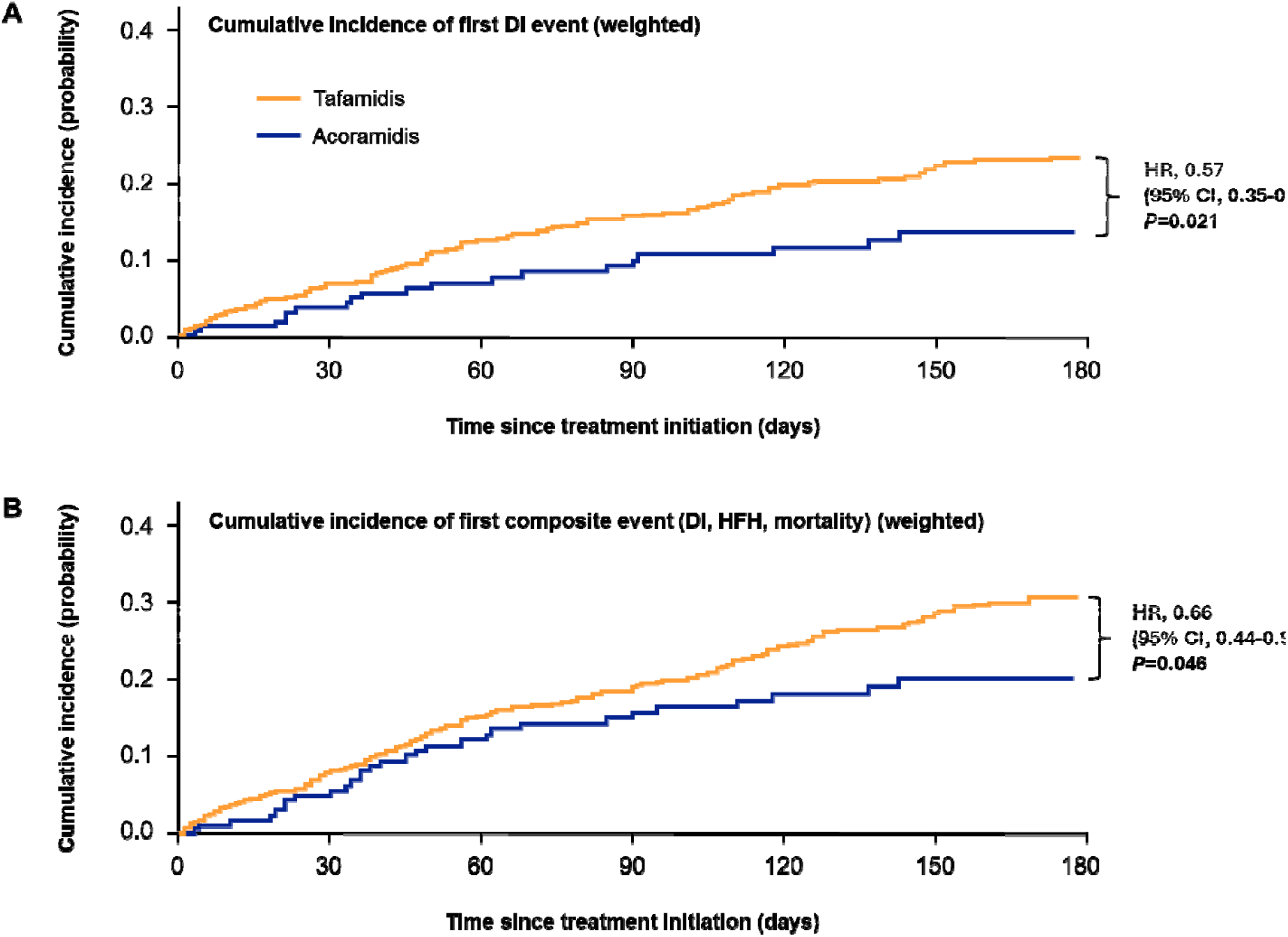
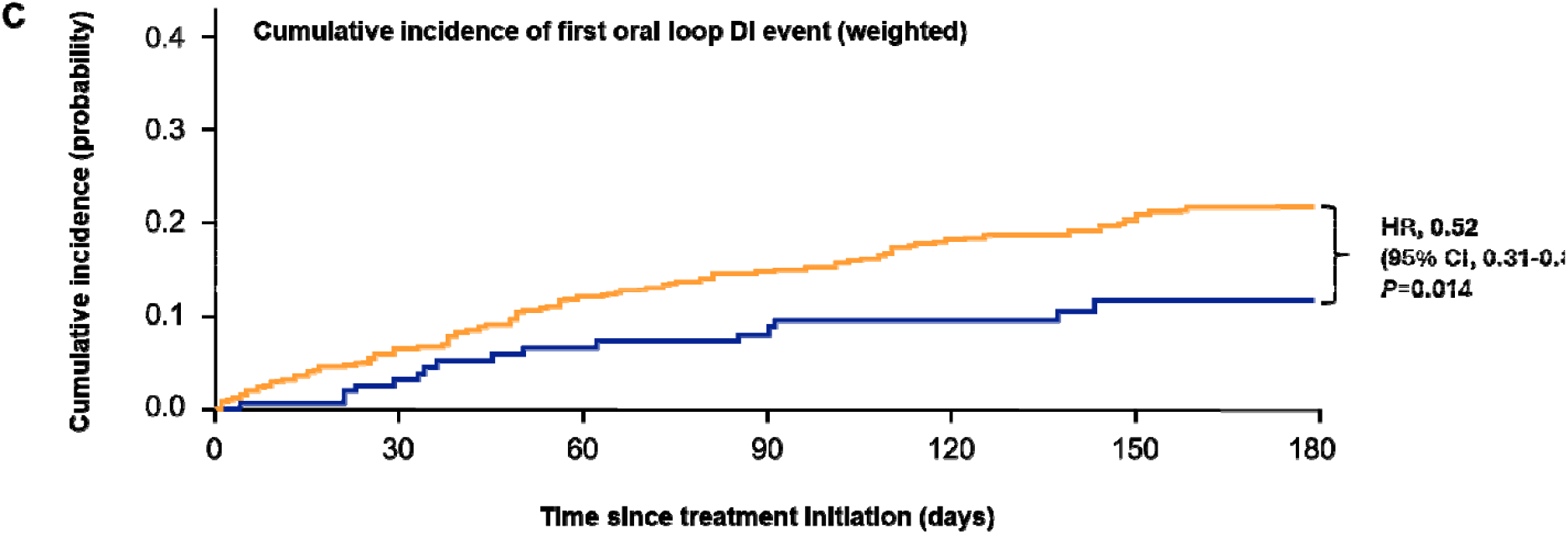
Cumulative Incidence of Real-World Outcomes. Real-world cumulative incidence of the first event of (A) DI; (B) the composite of DI, HFH, and mortality; and (C) oral loop DI among patients with ATTR-CM beginning at treatment initiation (weighted). The patient identification period was 12/11/2024 to 4/30/2025 and the study end date was 7/31/2025. DI, diuretic intensification; HFH, heart failure-related hospitalization; HR, hazard ratio.

## Discussion

In this first, robust real-world comparative effectiveness study of patients with ATTR-CM newly initiating a TTR stabilizer, acoramidis demonstrated a 43% and 34% reduction in risk of DI events and composite events (DI, HFH, mortality), respectively, compared with tafamidis. The cumulative incidence curves separated early and maintained separation over time.

Although definitions for DI may vary between studies,^4,6,9,10^ it is accepted that DI is associated with worse clinical outcomes.^11^ Monitoring outpatient oral loop DI is recommended every 12 months to determine disease progression.^4^ The larger reduction in DI events observed with acoramidis in the current study suggests early and improved clinical stability compared with tafamidis. The trend of reduction in HFH with acoramidis lends support to this interpretation, as patients with ATTR-CM are often hospitalized due to functional decline and recurrent instability, and hospitalization is associated with elevated mortality risk.^4^

Strengths of this study include the new-user active-comparator design, national claims dataset uniquely tokenized to hub data, prespecified clinically meaningful endpoints, inclusion of parenteral loop or oral thiazide-like diuretics for a comprehensive evaluation of DI, confounder adjustment via weighting, and use of multiple falsification outcomes that detected no bias. This real-world dataset includes higher proportions of under-represented populations, notably Black and female patients, than most ATTR-CM trials. This study is limited by missing clinical information not typically captured in claims data (eg, New York Heart Association class, N-terminal pro-B-type natriuretic peptide, genotype) and potential residual confounding despite weighting.

## Conclusions

In this first real-world comparative effectiveness study, acoramidis demonstrated a significantly lower risk of DI and a clinical composite of DI, HFH, and mortality compared with tafamidis. This may inform treatment selection for newly diagnosed patients and potentially support switching from tafamidis to acoramidis to prevent clinical events. Further study with longer follow-up and larger cohorts is warranted.

## Data Availability

Qualified academic investigators may submit requests for access to data via. Requests for access to study data will be evaluated by BridgeBio Pharma, Inc. and access will be provided contingent upon the approval of a research/study proposal and the execution of a data sharing agreement. BridgeBio Pharma will consider requests for access to participant-level data if participant privacy is assured through methods such as data deidentification, pseudonymization, or anonymization (as required by applicable law), and if such disclosures were included in the relevant studys informed consent form or study protocol.

## Data Sharing Statement

Qualified academic investigators may submit requests for access to data via. Requests for access to study data will be evaluated by BridgeBio Pharma, Inc. and access will be provided contingent upon the approval of a research/study proposal and the execution of a data sharing agreement. BridgeBio Pharma will consider requests for access to participant-level data if participant privacy is assured through methods such as data deidentification, pseudonymization, or anonymization (as required by applicable law), and if such disclosures were included in the relevant study’s informed consent form or study protocol.

## Author Contributions

All the authors participated in the conception and design of the study and in the analysis and interpretation of data. All the authors participated in the preparation and final approval of the manuscript.

